# Sulcal Patterns of the Medial Cerebral Cortex: A Comprehensive Scoping Review of Morphological and Morphometric Evidence

**DOI:** 10.1101/2025.09.17.25334987

**Authors:** Priyanka R Gohil, Priyanka N Sharma, Hetal V Vaishnani

**Affiliations:** Department of Anatomy, Smt. B K Shah Medical Institute and Research Centre, Sumandeep Vidyapeeth Deemed To be University, Piparia, Vadodara, Gujarat, India

**Keywords:** medial cerebral cortex, sulcal morphology, cadaveric anatomy, microneurosurgery, morphometry

## Abstract

**Background:** The medial cerebral cortex contains several sulci of high anatomical and clinical relevance, including the cingulate, paracingulate, calcarine, parieto-occipital, callosal, Rostral, supra-rostral and subparietal sulci. These structures serve as essential neuroanatomical landmarks and surgical corridors in microneurosurgical procedures, yet they exhibit considerable morphological variability. Although numerous studies have examined these sulci, no comprehensive synthesis exists focusing exclusively on cadaveric morphological and morphometric data, which remain critical for accurate neuroanatomical understanding.

**Methods:** This scoping review was conducted in accordance with the Joanna Briggs Institute (JBI) methodology and reported following the PRISMA-ScR checklist. A comprehensive literature search identified 4,440 records, of which 60 duplicates were removed. Screening of titles and abstracts excluded 3911 records, leaving 469 for full-text review. After applying eligibility criteria, eight cadaveric studies were included. Data were extracted on sample characteristics, morphological classification, and quantitative morphometry for the medial sulci. Findings were synthesized narratively and tabulated by sulcus type.

**Results:** The included studies analyzed a total of 422 hemispheres from formalin-fixed cadaveric brains. The cingulate sulcus was consistently present in all examined specimens, whereas the paracingulate sulcus displayed marked variability. The calcarine sulcus demonstrated relatively stable morphometry, with mean anterior and posterior segment lengths ranging from 2.3 to 3.5 cm, yet exhibited variable bifurcation patterns and lunate sulcus connections. The parieto-occipital sulcus was a reliable boundary between the cuneus and precuneus, with mean lengths around 4.0 cm. The subparietal sulcus was described less frequently, highlighting a gap in detailed morphometric literature.

**Conclusion:** Cadaveric evidence confirms both consistent and highly variable features in the medial cerebral sulci. These variations have direct implications for surgical planning, particularly in interhemispheric approaches. The paucity of detailed morphometric descriptions for certain sulci, especially the subparietal, callosal, rostral and supra-rostral sulcus, underscores the need for further targeted anatomical research.

## Introduction

The medial surface of the human cerebral cortex contains several prominent sulci that serve as fundamental landmarks for cortical organization and functional localization. The cingulate sulcus (CS) runs parallel to the corpus callosum and defines the superior border of the cingulate gyrus, a core limbic region involved in cognition and emotion [1–6]. The paracingulate sulcus (PCS), when present, lies dorsal to the CS and separates the medial superior frontal gyrus from the paracingulate gyrus [4,6–14]. Posteriorly, the parieto-occipital sulcus (POS) demarcates the boundary between the parietal and occipital lobes [15–21], while the calcarine sulcus (CalS) divides the occipital lobe into the cuneus and lingual gyrus and houses the primary visual cortex [22–27]. Collectively, these sulci provide essential anatomical landmarks used in neuroimaging, functional mapping, and neurosurgical planning.

Despite their clinical importance, considerable variability exists in the morphology and morphometry of these sulci. The PCS, in particular, exhibits striking hemispheric and sex differences: it is more often present and prominent in the left hemisphere, while frequently absent in the right [6,9]. Quantitative morphometric studies confirm that males often demonstrate greater left-sided fissurization, whereas females tend toward greater sulcal symmetry [6,20]. Such differences extend beyond normal variation; recent work indicates that reduced PCS length is associated with visual hallucinations in Parkinson’s disease [28] and with altered cognition in schizophrenia [3,8,11]. Similarly, the CS, POS, and CalS show individual variability in continuity, branching patterns, and depth, with implications for functional mapping and structural interpretation [1,29].

Cadaveric studies remain the gold standard for defining sulcal morphology, yet the literature is fragmented. Early anatomical reports were descriptive and based on small samples, whereas more recent neuroimaging studies often lack direct validation against dissection findings [6,29]. A systematic review of sulci and gyri morphology identified only a handful of cadaveric studies, most of which were methodologically limited and provided incomplete morphometric data [21]. No comprehensive synthesis exists that consolidates cadaveric evidence on the cingulate, paracingulate, rostral, callosal, parieto-occipital, and calcarine sulci. This absence of structured evidence mapping hinders the development of reliable anatomical reference standards.

This scoping review aims to systematically chart cadaveric evidence on the morphology and morphometry of the medial surface sulci-the cingulate, paracingulate, rostral, callosal, parieto-occipital, and calcarine sulci. Specifically, it will extract and synthesize quantitative and qualitative data, including sulcal length, depth, width, branching, laterality, and sex differences.

A consolidated understanding of sulcal variability holds direct clinical significance. Sulci serve as fundamental surgical corridors and orientation markers in neurosurgery [21], and their anatomical variability influences both functional localization and structural neuroimaging interpretation [1,28]. By mapping available cadaveric evidence, this review will provide a foundational anatomical reference, inform the creation of more accurate brain atlases, and guide future clinical and neuroimaging research.

## Methods

### Protocol and Reporting Framework

This scoping review strictly adhered to the Joanna Briggs Institute (JBI) methodological framework for scoping reviews and reported following the Preferred Reporting Items for Systematic Reviews and Meta-Analyses extension for Scoping Reviews (PRISMA-ScR) checklist. The review protocol was registered on the Open Science Framework: https://osf.io/9c5av [30]

### Eligibility Criteria

We included original cadaveric anatomical studies that described the morphology and/or morphometry of the cingulate, paracingulate, calcarine, parieto-occipital, or subparietal sulci in human brains. Both descriptive and quantitative studies were included. We excluded radiological studies, animal studies, case reports, review articles, conference abstracts without full text, and studies not published in English.

### Information Sources and Search Strategy

A comprehensive literature search was conducted in PubMed and Google Scholar from the database. The search strategy combined terms for each sulcus of interest with keywords related to cadaveric studies and neuroanatomy e.g. (“cingulate sulcus” OR “paracingulate sulcus” OR “calcarine sulcus” OR “parieto-occipital sulcus”) AND (“cadaveric study” OR “gross anatomy” OR “dissection”) AND (“morphometry” OR “morphological” OR “measurement” OR “depth” OR “length”). The reference lists of included studies were screened to identify additional eligible publications.

### Study Selection

The studies considered eligible for inclusion in this scoping review were human cadaveric studies evaluating the adult cadaveric brain specimens. Radiological studies of Paediatric and any clinical conditions, as well as studies in other species and articles without full text available, were excluded from this scoping review.

### Data Charting Process

Data extraction was performed independently by two reviewers using a standardized charting form. Extracted variables included: Author(s) and year of publication, Country of study, Sample size and number of hemispheres examined, Population characteristics (age, sex, laterality if available), Sulci examined, Morphological classifications, Morphometric measurements (length, branching patterns, distances to landmarks), Notable anatomical variations, and Key conclusions and clinical relevance.

### Data Synthesis

Findings were synthesized descriptively and grouped by sulcus type. Morphometric results were summarized in tables with ranges, means, and standard deviations where available. Morphological patterns were reported as frequencies or proportions. Due to the descriptive nature of the data and variability in measurement methods, no meta-analysis was performed.

## Results

A total of 4,440 records were retrieved, of which 60 duplicates were removed. After title and abstract screening, 3,911 records were excluded. Full-text review was performed for 469 articles, resulting in the exclusion of 461 papers for reasons including non-cadaveric methodology, pediatric or pathological specimens, non-medial sulci, and lack of full text. Ultimately, eight cadaveric studies met inclusion criteria, collectively examining 422 hemispheres. The study selection process is summarized in Figure 1.

**Figure 1.**
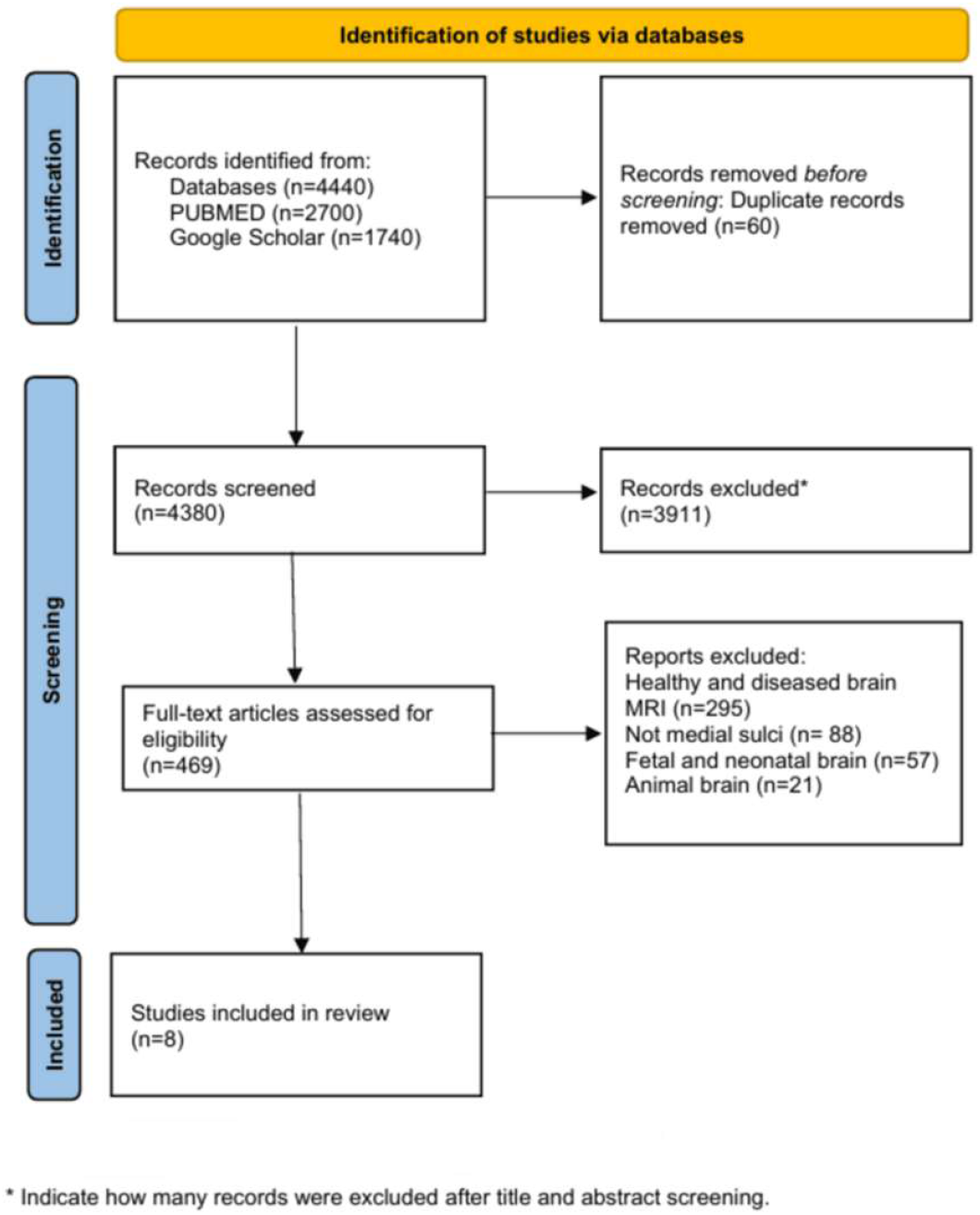
Study selection process (PRISMA-ScR Checklist)

The included studies were conducted in diverse populations (European, Indian, Japanese). However, demographic information such as age and sex was incompletely reported, limiting subgroup analysis. The findings are organized below by sulcus.

### Cingulate and Paracingulate Sulci

There is a lack of specific morphometric and morphological data on cadaveric studies reporting the average length of the cingulate sulcus in adult cadavers. Selahi et al. stated that the PCS was present/prominent in roughly 25% of specimens, whereas the intralimbic sulcus was seen in about 15%. These dissection results align with the MRI findings that PCS prominence is a minority variant and support observed sex differences (males showed more prominent PCS)[12]. Imada et al. (2021) performed detailed morphological observations and classified the medial frontal cortex (excluding the cingulate gyrus) into 2-4 gyri, with 56.6% of hemispheres showing 3 gyri [31]. These sulcal and gyral arrangements were considered reliable intraoperative landmarks for anterior interhemispheric approaches.

### Calcarine Sulcus (CalS)

Mandal et al. (2014) examined 106 cadaveric brain hemispheres and found bifurcation of the calcarine sulcus into two rami in 59.43% of specimens. Direct continuation to the lunate sulcus occurred in 31.13% of cases, with notable morphological variations including “S” and “f” shaped terminations [32]. Chandra et al. (2018) reported mean lengths of the anterior and posterior CalS segments as 2.3 ± 0.43 cm and 3.5 ± 0.38 cm, respectively and Morphometric differences between right and left hemispheres were noted, but were statistically not significant (n=100 hemispheres) [27]. Malikovic et al. (2012) classified four morphological types: Type I (single apex), Type II (two apexes), Type III (S-shaped), and Type IV (horizontal)[33]. Approximately 80% of hemispheres demonstrated posterior bifurcation into upper and lower branches (n=30 hemispheres). In 13.3%, the posterior CalS was separated from the middle portion by a cuneolingual gyrus. These patterns are directly relevant to the localization of the primary visual area (V1).

### Parieto-Occipital Sulcus (POS)

The POS originated from the CalS and ran upwards and posteriorly towards the superomedial border. Nayak et al. (2023) described the POS as originating from the CalS and extending toward the superomedial border in which the mean distance from CalS bifurcation to POS termination was 4.08 cm (n=31 hemispheres, 17 right and 14 left with unknown sex) [34]. Table:1 shows quantitative evidence of length of sulci present on medial cerebral cortex.

**Table 1.**
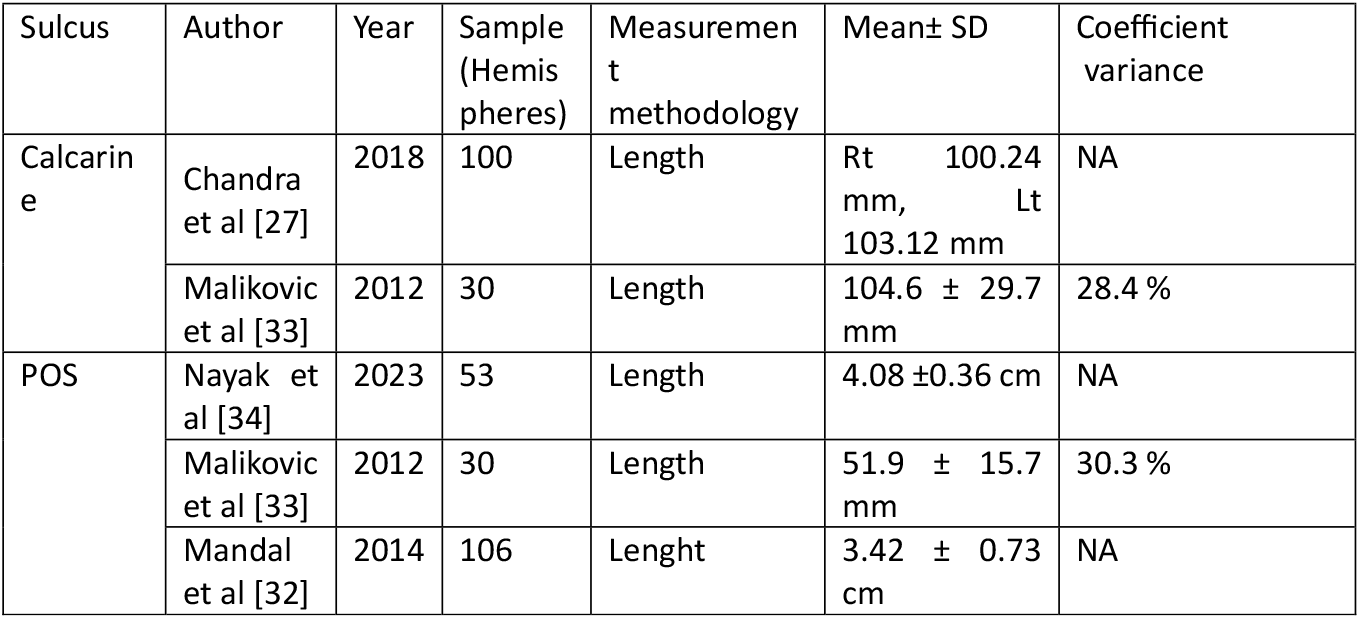
Morphometric Evidence on sulci presents on medial surface.

| Sulcus | Author | Year | Sample (Hemispheres) | Measurement methodology | Mean± SD | Coefficient variance |
| --- | --- | --- | --- | --- | --- | --- |
| Calcarine | Chandra et al [27] | 2018 | 100 | Length | Rt 100.24 mm, Lt 103.12 mm | NA |
|  | Malikovic et al [33] | 2012 | 30 | Length | 104.6 ± 29.7 mm | 28.4 % |
| POS | Nayak et al [34] | 2023 | 53 | Length | 4.08 ±0.36 cm | NA |
|  | Malikovic et al [33] | 2012 | 30 | Length | 51.9 ± 15.7 mm | 30.3 % |
|  | Mandal et al [32] | 2014 | 106 | Length | 3.42 ± 0.73 cm | NA |

Malikovic et al. (2012) identified three morphotypes: Type I (straight, unbranched), Type II (Y-shaped, more frequent in the left hemisphere), and Type III (T-shaped with horizontal superolateral branch, longest in right hemisphere). Across studies, the POS reliably demarcated the cuneus from the precuneus, serving as a robust anatomical landmark for visual cortical mapping. POS consistently connects with the calcarine sulcus at a junction known as the cuneal point, serving as a reliable anatomical landmark for mapping visual cortical areas. While its length varies slightly between hemispheres, type 3 tends to be the longest, and type 1 was the shortest [34]. Table:2 provides detailed summary of morphologicalevidences.

**Table 2.** Morphological Evidences of sulci present on medial surface.

| Sulcus | Authors | Year | Demography | Sample (Hemispheres) | Morphological findings | Variability |
| --- | --- | --- | --- | --- | --- | --- |
| Cingulate | Selahi et al [12] | 2022 | Turkey | 20 | Present in all cases. | Associated with intralimbic sulcus (15%) |
| Paracingulate | Selahi et al [12] | 2022 | Turkey | 20 | Prominent (10%), Present (15%), Absent (75%) | Leftward asymmetry, more prominent in males. |
| Calcarine | Mandal et al [32] | 2014 | India | 106 | Bifurcation in (59.43 %), Continuation to lunate sulcus (31.13%) | “S” and “F” shaped terminations |
| Parieto-occipital | Nayak et al [34] | 2023 | India | 31 | Originates from Calcarine sulcus; distinct boundary between cuneus & precuneus | Morphology consistent |
| Subparietal | Gurer et al [17] | 2013 | USA | 56 | Consistent location relative to marginal ramus and POS | Lacks morphometric details |

### Subparietal, Callosal, and Rostral sulci

The subparietal sulcus was consistently located relative to the marginal ramus of the cingulate sulcus and the POS [17], but no detailed morphometric data were available. The callosal and rostral sulci were rarely described in cadaveric studies; available reports provided only qualitative anatomical description without measurements. Overall, quantitative data on these sulci remain a major gap in cadaveric neuroanatomical literature.

## Discussion

This scoping review synthesizes cadaveric morphological and morphometric evidence for the medial cerebral sulci-specifically the cingulate, paracingulate, calcarine, parieto-occipital, Rostral and subparietal sulci-based on eight anatomical studies published between 1995 and 2025. By consolidating these findings, we highlight consistent sulcal patterns that serve as reliable intraoperative landmarks, as well as notable anatomical variations with potential surgical implications.

### Morphological Consistency and Variability

The cingulate sulcus emerged as a highly consistent structure, present in all examined hemispheres across included studies. In contrast, the paracingulate sulcus demonstrated substantial variability, with absence in up to 75% of hemispheres, and a tendency for greater prominence on the left, particularly in males. This asymmetry has been observed in neuroimaging literature and may influence approaches to the medial frontal lobe, where identification of a prominent paracingulate sulcus can alter surgical corridors [12].

Calcarine sulcus morphology exhibited less variation in presence but notable variability in branching patterns and terminal morphology. Mandal et al. (2014) documented bifurcation into two rami in over half of specimens and reported unusual terminations such as “S” and “f” shapes, which may pose challenges for surgical localization of the primary visual cortex [33].

The parieto-occipital sulcus (POS) consistently served as a reliable boundary between the cuneus and precuneus, with minimal morphological variation and this reliability enhances its utility as a landmark in posterior interhemispheric approaches [34]. In contrast, the subparietal sulcus remains underdescribed in cadaveric literature, despite its relevance to the posterior cingulate and retrosplenial cortex.

### Comparison With Previous Literature

Radiological studies have also reported variability in paracingulate sulcus prevalence and morphology, often citing rates of 30-60% [1,6,21]. Our cadaveric synthesis shows a lower prevalence of prominent paracingulate sulci, which may reflect population-specific differences or methodological variation. Similarly, while imaging-based morphometry of the CalS often reports more symmetric lengths between hemispheres, cadaveric evidence confirms small but measurable asymmetries [24]. In neurosurgery, accurate identification of medial sulci is essential for safe entry into interhemispheric fissures and deep cortical structures. The cingulate and paracingulate sulci guide access to the anterior cingulate gyrus and supplementary motor area, regions implicated in tumor resections and epilepsy surgery [32]. Variability in paracingulate sulcus presence necessitates careful preoperative imaging correlation to avoid misinterpretation of cortical boundaries. The calcarine and parieto-occipital sulci are critical for visual pathway preservation. Misidentification of CalS bifurcation or POS origin during posterior approaches could increase the risk of postoperative visual field deficits. Morphometric data from cadaveric studies provide intraoperative measurement benchmarks when sulci are obscured by pathology or distortion. The relative lack of quantitative data for the subparietal sulcus suggests an important avenue for future anatomical research. Given its proximity to the posterior cingulate cortex-a target in functional neurosurgery-detailed morphometric characterization is warranted.

### Limitations

The included studies varied in sample size, population demographics, and measurement techniques, limiting direct quantitative synthesis. Most lacked comprehensive demographic data, making it difficult to assess sex-or age-related variation. Furthermore, the predominance of studies from single geographic regions may reduce generalizability.

## Data Availability

All data produced are available online at https://osf.io/y8ec7/

